# Continuous Glucose Monitoring in Older In-patients with Type 2 Diabetes and Cognitive Impairment – An open single-arm feasibility study

**DOI:** 10.64898/2026.01.26.26344013

**Authors:** Busra Donat Ergin, Katharina Mattishent, Anne Marie Minihane, Richard Ian Gregory Holt, Helen Murphy, Ketan Dhatariya, Michael Hornberger

## Abstract

**Background:** Type 2 diabetes (T2DM) and cognitive impairment are common long-term chronic conditions affecting older people in hospital. Cognitive impairment can complicate glucose monitoring and lead to diabetes-related emergencies in T2DM. Traditionally, point of care test (POCT) measurements of capillary blood glucose are conducted in-hospital for T2DM while continuous glucose monitoring (CGM) is not widely used.

**Aim:** To understand the feasibility, acceptability and tolerability of using CGM in older in-patients with T2DM and cognitive impairment.

**Methods:** Thirty-two older people (mean age = 78.7 ±6.7 years) with comorbid T2DM and cognitive impairment (AMT ≤8/10 and Mini-ACE ≤22/30) were recruited within a tertiary care hospital in the UK. All participants were naive to CGM and were asked to wear blinded Dexcom G7 sensors for up to 10 days. Participants were asked about feasibility, acceptability and tolerability questions at the point of sensor removal.

**Results:** Twenty-nine participants (96%) reported no pain during CGM fitting. All participants (100%) agreed that they did not notice wearing the sensor, and it did not affect their day-to-day hospital activities. All participants (100%) found it ‘very easy’ or ‘easy’ to have the sensor fitted and wearing it for 10 days, with 27 participants (90%) finding CGM convenient. Seventeen participants (57%) reported favourable perceptions of the subcutaneous sensor sensation.

**Conclusion:** CGM use in older in-patients with T2DM and cognitive impairment is highly feasible and acceptable for patients. Future studies and trials are now needed to evaluate the clinical use of CGM for glucose monitoring in hospitalised or community-dwelling older individuals with T2DM and cognitive impairment.

## Introduction

Type 2 diabetes (T2DM) is a common long-term condition associated with a loss of insulin sensitivity and hyperglycaemia mostly affecting adults, particularly older adults. Individuals aged 65 years and older with T2DM constitute nearly half of the total population of people living with diabetes [1].

Chronic complications of diabetes increase with the duration of disease and can lead to disability, lower quality of life and eventually higher mortality. While the most common chronic complications are cardiovascular diseases, gangrene and stroke [2], one of the long-term common complications of diabetes is dementia [3]. Diabetes significantly increases dementia risk, particularly with longer diabetes duration or presence of hypoglycaemia [4]. This cognitive impairment emerges through complex interactions involving insulin resistance, brain hypometabolism, inflammatory processes, vascular injury, and reduced cerebral blood flow, and oxidative stress [5].

Cognitive impairment can complicate diabetes self-management, including glucose monitoring, further exacerbating the risk of diabetes complications and dementia pathophysiology progression. Advances in technology for better diabetes management such as continuous glucose monitoring (CGM) can contribute to decreasing the number of complications as well as the risk of cognitive decline [2]. Recent data shows that holistic and individualised approach is the ideal pathway for diabetes treatment for older adults with a variety of comorbidities with T2DM and other comorbidities [6]. CGM is emerging as a valuable technology due to its safety and practicality, particularly for individuals with multimorbidity or geriatric syndromes such as cognitive impairment [7]. However, more research is needed to understand the benefits and challenges of CGM in this population with T2DM and cognitive impairment.

Traditionally, point of care test (POCT) measurements of capillary blood glucose are conducted in-hospital for T2DM whilst continuous glucose monitoring (CGM) is not currently used in the wards since there is no established feasibility of using such devices within the hospital[8]. This feasibility study aims to investigate the feasibility, acceptability and tolerability of CGM in older adults with T2DM and cognitive impairment while being in-patients.

## Methodology

### Study Design

A mixed-method open-label single-arm observational feasibility study of CGM among older people with T2DM and cognitive impairment who are in-hospital patients.

### Participants: Inclusion and Exclusion Criteria

Eligible participants were older (≥65 years) in-patients with T2DM for at least 6 months and who had been treated with anti-diabetes drugs (either oral, non-insulin injectables or insulin) (i.e. not managed with diet and exercise alone).

Participants were also required to have an Abbreviated Mini-Mental Test (AMT) score equal or less than 8 (out of 10) and/ or a Mini-Addenbrooke’s Cognitive Examination (Mini-ACE) UK Version A score ≤21 (out of 30), indicating any level of cognitive impairment, provided they retained decision-making capacity. The AMT is commonly used, brief cognitive screening tool within hospitals in the UK. The Mini-ACE is a cognitive screening tool sensitive to earlier cognitive changes using two cut-offs (25 or 21 out of 30) [9], and has been validated in various settings.

Potential participants were excluded if they did not have mental capacity and/or were on end-of-life care, as confirmed by the clinical care teams. We also excluded people who were current CGM users, were undergoing dialysis treatment and showed evidence of skin irritations on the upper arm or abdomen where the CGM sensor would potentially be fitted. We also excluded people who had other neurological or psychiatric diseases, as well as those with autism, dyslexia or learning difficulties because these conditions could confound cognitive test results.

### Setting

The study was conducted in a tertiary care hospital in England (Norfolk and Norwich University Hospitals NHS Foundation Trust) and the CGM sensor was fitted whilst the consented participants were in-patients (as detailed in the recruitment process section).

### Intervention

The Dexcom G7 CGM device is licensed for use in children and adults and available for consumers to directly purchase. The kits compromise auto applicator kits with sensors with transmitter and receivers to be used in the blind mode. We provided blinded receivers to all participants which was paired with their sensors in advance.

The sensors were applied to the back of the upper arm. Interstitial glucose readings were measured continuously, and data was transmitted from the sensor every 5 minutes. Time in range (TIR) was defined as the time spent when the glucose concentration was between 4 mmol/L to 10 mmol/L in line with international recommendations and default setting [10], [11], [12].

### Recruitment process

One of the study members investigated the initial selection of potential participants via the hospital electronical system (Integrated Clinical Environment: ICE) by checking admissions according to the inclusion criteria, except eligibility criteria of AMT and Mini-ACE, and approached the potential participants with a consent to contact form. We prioritised geriatric wards and wards where patients were expected to have longer hospital stays.

Potential participants were subsequently given the participant information sheet (PIS) and allowed 24 hours to consider taking part in the study. After 24 hours, potential participants were revisited, and if they agreed to participate, participants provided written consent. No incentives were offered for participation. After consent, participants were assessed for cognitive impairment via the AMT and Mini-ACE. If participants had a score of 8 or lower on the AMT and/or a score of 22 or lower on the Mini-ACE, they were invited to take part in the study. If the cognitive screening scores indicated no cognitive impairment, participants were told that they were not eligible for participation. Figure 1 provides a detailed CONSORT diagram of the recruitment process.

Participants continued to receive their usual hospital care and with their usual diabetes management during the study period.

### Baseline assessment

A research team member fitted a CGM sensor on the participant’s upper arm and blinded the receiver. Participants sociodemographic details (sex, age, ethnicity, marital status, smoking and driving status) were obtained.

### Follow-up assessment

The follow-up assessment was conducted on the 10^th^ day after CGM sensor fitting. The 10-day study period was chosen due to the manufacturers recommendation to change the sensor after 10 days and considering that some people would not be hospitalised for longer than 10 days. The participant was visited at their hospital bed or if they had been discharged, at their residence (private or care home). If the participants were discharged within 10 days, they were discharged wearing the sensor and taking the receiver, before a research team member removed the sensor on the 10^th^ day and uploaded the data to Dexcom Clarity^®^ system anonymously. The participants were asked CGM feasibility, acceptability and tolerability questions via a questionnaire, including open-ended questions. Finally, participants underwent the same cognitive testing as during the first research appointment and a further DBS sample was collected. Clinical data for all participants was collected via the hospital’s electronical clinical care management system. Figure 2 demonstrates the study design and procedures briefly.

### CGM feasibility, acceptability and tolerability survey

Feasibility, acceptability and tolerability were determined via a questionnaire (Appendix), including 1 pain scoring question, 2 dichotomous, 7 Likert-style multiple choice questions, and 2 open-ended questions. We encouraged participants to talk about their experiences and additional thoughts about the device with open-ended questions. The questionnaire was modified from a previous CGM feasibility study [13].

### Statistical analysis plan

All statistical analyses were conducted using RStudio (R version 4.3.2; R Foundation for Statistical Computing, Vienna, Austria). Given the feasibility nature of the study, analyses were primarily descriptive and exploratory. Descriptive statistics were used to summarise feasibility, acceptability, and tolerability survey outcomes, participant characteristics, clinical data and glucose-related metrics. Continuous variables were reported as mean with standard deviation (SD) or median, while categorical variables were summarised using frequencies and percentages.

Comparisons between CGM-derived glucose metrics and POCT-derived measurements were performed to assess differences between measurement methods. Pre- and post-CGM cognitive scores for AMT and Mini-ACE were compared using paired t-tests. Statistical significance was defined as a two-tailed p-value < 0.05.

### Sample size calculation

The sample size was based on existing feasibility studies within the field [13], [14], as well as a prior hospital audit of inpatient data meeting the study criteria.

### Ethics

This study has been reviewed and approved by Health Research Authority (HRA) and Health and Care Research Wales (HCRW) on 23^rd^ August 2024 (IRAS project ID: 344263, REC reference: 24/LO/0441).

### Patient and Public Involvement and Engagement (PPIE)

Patient and public involvement was conducted during study development process. Two individuals over 65 years with type 1 diabetes and prior CGM experience provided feedback on study design and acceptability through a consultation meeting, which informed the study planning.

## Results

A total of 32 participants were recruited, with the final sample comprising 30 participants. We excluded 2 participants: one due to more than 50% missing CGM readings during the 10-day study period; and the second because they were not taking antidiabetic medication.

### 1. Sociodemographic Characteristics

The mean age was 78.8 ± 6.7 years, with 18 male (60%) and 12 female (40%) participants. 13 participants (43.3%) were living alone, whilst the remainder were living with a partner, child, grandchildren (n=15, 30%) or in a care home (n=2, 6.6%). All participants were retired. Two thirds were secondary school graduates, with 6 (20%) having a higher education degree (Bachelor’s or higher) and 5 (16.6%) did not finish high school. We summarized main characteristics of the participants in Table-1.

**Table-1:** Participant Characteristics.

| Participant characteristics | Mean | Median | Min | Max |
| --- | --- | --- | --- | --- |
| Age (years) ± SD | $78.7 \pm 6.7$ | 78 | 67 | 94 |
| BMI ($\text{kg}/\text{cm}^2$) ± SD | $31.2 \pm 10.9$ | 28.3 | 20.7 | 74.3 |
| HbA1c ($\text{mmol}/\text{mol}$) ± SD | $75.1 \pm 29.5$ | 67 | 45 | 154 |
| Medications (N) | $10.5 \pm 4.4$ | 11 | 3 | 20 |
| Hospitalisations (N) in the last 12 months (excluding current admission) | $0.8 \pm 1.5$ | 1 | 0 | 8 |
| Length of T2DM (Years) | $14.9 \pm 6.4$ | 12.5 | 0.6 | 38 |

Only 5 participants (16.6%) were walking for pleasure or shopping with 3 out of the 5 walking less than 1 hour per week. Seven participants (23.3%) were doing light housework such as cooking and cleaning for less than 3 hours/week and eight (26.6%) were doing gardening or do-it-yourself (DIY) activities. 43.3% were sedentary; however, ten participants had at least one leg ulcer, lower limb cellulitis or diabetic foot, and two had leg amputation which affected physical activity levels. The mean frailty score among our participants was 5.83 ± 0.83 (using 1-9 scale, fitness vs. vulnerability), which indicates that the majority of them were mildly frail.

### 2. Clinical Data

Six (30%) were admitted as in-patients due to diabetes related issues (e.g. DKA, hypoglycaemia, hyperosmolar hyperglycaemic state (HHS). Another third was admitted due to a fall, either due to hypoglycemic or other reasons. One was admitted due to cognitive impairment and confusion. Twenty-four (80%) participants had at least four comorbidities at admission.

One participant had a diagnosis of dementia, which was Lewy body dementia, for 1 year. At the second research appointment, fourteen participants (46.6%) were still in hospital, including two (6.6%) at a different hospital for rehabilitation. Two participants (6.6%) were discharged to care home or nursing home, while fourteen (46.6%) were at their private home at the end of the study.

Thirty percent of participants were treated with insulin, while 46.6% were on oral glucose anti-diabetes agents, and 30% of the participants were using both insulin and oral agents (Table-2). Due to diabetic foot or foot ulcers, 56.6% were regularly using topical agents. Supplements and multivitamins (mostly vitamin D, folic acid and thiamine) were commonly used (73.3%).

**Table-2:** Medication classes.

| **Anti-Diabetic medications\*,\*\*** | **Total Number of participants N (%)** |
| --- | --- |
| **Insulin (solely)** | 7 (23.3%) |
| **Insulin and oral antidiabetics (combined)** | 10 (33.3%) |
| **Metformin** | 16 (53.3%) |
| **SGLT-2 inhibitors** | 9 (30%) |
| **DPP4 inhibitors** | 4 (13.3%) |
| **Sulphonylurea** | 3 (10%) |
| **GLP-1 receptor agonists** | 1 (3.3%) |
| **Commonly prescribed other medications\*,\*\*** |  |
| **Statins** | 20 (66.6%) |
| **Anticoagulants/Antiplatelets** | 20 (66.6%) |
| **Topical agents** | 17 (56.6%) |
| **Analgesics** | 15 (50%) |
| **Antibiotics** | 8 (26.6%) |

*\*The medications are the regular medications of our participants recorded at admission.*
*\*\*There was overlap between the groups; for example, some metformin users were also prescribed DPP4 inhibitors, and some inpatients receiving statins were also taking anticoagulants.*

### 3. Feasibility, Acceptability and Tolerability Outcomes Feasibility

All participants (100%) found it ‘very easy’ or ‘easy’ to have the sensor fitted and keeping CGM for 10 days.

#### Acceptability

All participants (100%) reported they did not feel the sensor physically. Twenty-nine participants (96%) reported no pain during CGM fitting. All participants (100%) agreed that they were unaware of the sensor once fitted, and it did not affect their day-to-day hospital activities. All participants (100%) reported CGM did not make any difference to their symptoms or experiences of living with diabetes, any other conditions or self-care. Twenty-eight (93.3%) felt confident to wear CGM. Twenty-seven (90%) participants reported they were likely to recommend CGM to their friends or family.

#### Tolerability

Seventeen participants (56.6%) reported that positive sensor skin sensation, while the others reported they did not notice the existence of CGM. Twenty-seven participants (90%) found CGM convenient. Twenty-four (80%) reported, overall, they liked using CGM.

We summarised some of the feasibility, acceptability and tolerability feedback on CGM experience from our participants with Chart 1.

In our open-ended questions, most participants reported that they forgot the sensor’s existence and did not feel anything, and it was easy to keep CGM for 10 days (≥73.3%). Two participants mentioned unsolicited that they would prefer to use CGM compared to POCT in their daily life since they had sore fingers and numbness and found it painful to prick their fingers multiple times a day. Three participants (10%) reported that they needed to remind healthcare staff that they were wearing a CGM sensor, when they were measuring blood pressure via upper arm blood pressure cuffs. One participant (3.3%) reported that the sensor came off on the 4^th^ day, and that they did not wish to have a replacement sensor.

Three participants (10%) said they were happy to contribute to the research, as diabetes and dementia are very serious diseases and research in this field should be encouraged. One participant who had low nocturnal sensor reading commented “*It was good to see hypo readings during the night; I was not aware that I am having it frequently. But I knew it decreases sometimes. I will inform my diabetes nurse and GP about it and discuss CGM, my current medications and night-time snacks”*.

### 4. CGM vs POCT Readings

Mean time in range (TIR= 4-10 mmol/mol) was 36.2%, time above range (TAR ≥ 10 mmol/mol) was 62.8% and time below range (TBR ≤ 3.9 mmol/mol) was 1%. Fourteen (46.6%) participants wore their CGM continuously in hospital, with CGM active for 89.9% of the time (range 86% - 95%). Mean TIR based on available POCT readings was 40.8%, TAR was 57.2% and TBR 1.8%, showing similar readings as CGM-derived glucose metrics. However, there were individual differences between CGM and POCT, with some individuals showing no TBR (0%) via POCT, whilst CGM observed results 3% and 8% TBR within the same participants.

Overall, CGM data was very similar and consistent to POCT glucose measurements. One participant (3.3%) had an apparent CGM sensor fault at day 6 after sensor fitting, leading to consistent above range readings compared to POCT. For this participant, we only included CGM data up to day 6.

### 5. Cognitive Status

The participants were screened with AMT and Mini-ACE to decide their eligibility for this study. Then, all participants completed other cognitive tests after fitting CGM in the baseline assessment The mean AMT of the participants was 7.2 ± 2.2 and the mean Mini-ACE was 15.6 ± 4.1 (Table-3). We also compared AMT and Mini-ACE scores before and after using CGM using paired t-test analyses (n=28). There was no significant change in AMT scores (p=0.28), while Mini-ACE scores increased significantly (p<0.01), indicating cognitive improvement. Results of our study demonstrate how cognitive impairment common in this frail population, despite the absence of a formal dementia diagnosis.

**Table-3:** Cognitive Assessment.

| Cognitive Tests | Mean | Min | Max |
| --- | --- | --- | --- |
| AMT (baseline assessment) ± SD | $7.2 \pm 2.2$ | 3 | 10 |
| AMT (follow up assessment*) ± SD | $7.5 \pm 2.1$ | 3 | 10 |
| Mini-ACE (baseline assessment) ± SD | $15.6 \pm 4.1$ | 5 | 22 |
| Mini-ACE (follow up assessment*) ± SD | $18.5 \pm 4.9$ | 8 | 26 |

*\*2 participants (6.6%) did not complete the cognitive testing at the follow up appointment; 1 participant reported they were feeling unwell due to diabetic foot pain and did not wish to do cognitive tests, whilst the other participant was transferred from their ward to critical care unit.*

Overall, the results showed high feasibility and acceptability among in-patients with T2DM and cognitive impairment.

## Discussion

Our study suggests that CGM use is highly feasible and acceptable for older hospitalised individuals with T2DM and cognitive impairment. Specifically, the results of our study show that wearing CGM devices is highly acceptable, feasible and tolerable, even for people with significant cognitive impairment.

Specifically, our findings replicate a previous CGM feasibility study in the same population within the community, i.e. not in-hospital [13]. Both their and our findings demonstrate high feasibility and acceptability of CGM. In more detail, the feasibility was evident for all our participants, with no exception. Similarly, all acceptability scores were above 90% indicating that people often did not even notice wearing the CGM device after some time. Tolerability scores were slightly lower but still very high (90%) with participants indicating that they found CGM very convenient to use. Some people were encouraged to use CGM after their discharge from hospital. Further, some participants highlighted that they liked the detection of potential hypoglycaemic events, in particular at night, which would allow them to adjust their night-time snacks. Taken together, our findings demonstrated that CGM is highly acceptable, feasible and well tolerated by participants for people with comorbid dementia and cognitive impairment in-hospital.

The in-hospital setting was in itself challenging, with some potential participants finding it overwhelming to take part in our study part despite expressing initial interest (see Figure 1). It highlights the challenges of conducting research in inpatient settings. Yet, final feedback from consented participants showed that CGM did not affect day-to-day activities during their hospital stay, further highlighting that CGM is highly feasible within in this setting but that the setting itself was the main challenge. For this reason, larger prospective studies or clinical trials are warranted to evaluate the impact of CGM within this setting and to determine the optimal clinical implementation strategies for CGM use in this vulnerable inpatient group. In this regard, it is also interesting that half of our participants considered using CGM in the future, i.e. after discharge from the hospital. This highlights another research gap, in that people within the community could benefit from CGM use. Indeed, retrospective data analyses have suggested that CGM use in T2DM and cognitive impairment in the community can affect overall mortality and hospitalisation rates [15], [16]. One is RELIEF study showing that two years use of flash glucose monitoring was associated with 49% reduced hospitalization due to acute diabetic events mainly diabetic ketoacidosis (DKA) [17]. Future prospective CGM studies in the community settings in this regard are needed urgently to reduce mortality and hospitalisation in the first place of this vulnerable group.

In terms of the actual CGM readings, to which participants and hospital staff were blinded, the readings showed similar glucose level findings. While the TIR goal for glucose level between 70 and 180 mg/dL (3.9-10 mml/L) is >70% for the general population [18], consensus suggests less stringent thresholds for older people, with a TIR target of >50% [19]. Despite this lower TIR goal, the mean TIR in our population was still low. Lifestyle interventions such as regular physical activity, healthy and balanced diet and modest weight loss, have been recommended to improve glucose targets, however, our sample was clearly physically inactive, and many had severe diabetes related complications, including amputations, hindering their physical activity levels and increasing their frailty [20]. Further, multiple other reasons, including polypharmacy, perioperative procedures, and other hospital changes might affect the glucose readings of patients. Finally, clinical guidelines are clearly geared towards the reduction of hypoglycaemia [20], [21], which might explain the low TIR within our participants.

The CGM readings also detected 28 hypoglycaemic events (<=3.9 mmol/L) for 9 participants. Eleven of the hypoglycaemic events (39.3%) were nocturnal (12am-6am). However, low nocturnal sensor glucose readings in our study may not directly reflect true nocturnal hypoglycaemia and should be interpreted in light of potential sensor bias and sensor–clinical discordance, see also [22]. Previous studies suggests that CGM may show less accuracy with lower glucose ranges compared to POCT and highly recommend comparison to capillary glucose levels when possible [8], [23], [24], [25]. It highlights still the challenges using CGM in settings where people might have low readings that might not be ‘real’ or pose an immediate adverse risk within hospital settings.

In terms of the cognitive screening findings, our results showed that a discrepancy between the employed cognitive measures (AMT vs Mini-ACE). The AMT is widely used to assess cognitive status in hospital settings in the UK [26]. Although it has practical use on the wards and at hospital admission, there are some limitations regarding language and culture, as well as unreliability of certain questions and reduce sensitivity to detect cognitive changes cross sectionally and longitudinally [27], [28]. Because of these limitations, we included the Mini-ACE in our cognitive screening, which is commonly used in UK memory clinics for dementia screening and provides a quick cognitive screening read-out, similar to the MoCA (Note the Mini-ACE has been validated against the MoCA). Our results showed that the Mini-ACE is more sensitive and less likely to have ceiling effects than the AMT. We only applied the lower Mini-ACE cut-off (<=21/30), since this score is accepted as almost certain score for dementia or a person with cognitive impairment [29]. AMT results of potential participants were higher than Mini-ACE and would have resulted in exclusion of most potential participants. It suggests that the AMT might be not suitable to cognitively screen in hospital, as it only detects severe cognitive deficits, e.g. delirium, while potentially missing patients with mild to moderate cognitive deficits. There is a dearth of research on how cognitive impairment levels impact glucose management in older T2DM people. Our results show that despite all participants having mental capacity to consent to the research, there cognitive performance moderate cognitive impairment, further complicating diabetes management.

Interestingly, the cognitive scores for the AMT did not change from baseline to follow-up assessment (10 days later), whereas those for the Mini-ACE did. The Mini-ACE scores at the follow-up assessment were significantly higher than the baseline assessment scores. Since short follow-up duration and use of masked CGM are unlikely to provide meaningful cognitive changes, this might have been due to patients having recovered from their conditions, they were admitted for in the first place. It also highlights that cognition levels can change during hospital stays. In this regard, it was also telling that the AMT scores did not change from baseline to follow-up, further underlining that the AMT has limited sensitivity to measure mild to moderate cognitive impairment within hospital cross-sectionally and longitudinally. Future research on cognitive screening in hospital settings is needed to determine whether better tools than the AMT can be applied, as they might improve patient care, including glucose management.

Despite the novelty of our research with promising findings, there are some limitations to our study. Overall, this is a small pilot study in one hospital, which requires replication in a larger sample and different hospital settings. Further, participants were admitted to the hospital due to different health conditions including emergencies. Therefore, they were not in their best health, and this might have affected the cognitive and CGM results. We ran our recruitment search in particular wards due to our inclusion criteria (older age), and having longer hospital stays. This selection may have caused a potential bias in the recruitment process; for example, we recruited five participants from a general surgery ward which potentially resulted in lower physical activity scores in our study population since some participants were awaiting lower limb amputations. Despite this limitation, our findings were highly consistent regardless of health condition or ward recruited from. Finally, in our study, we used masked CGM, and it is, therefore, not clear whether CGM use can actually improve glucose management in hospital, compared to traditional POCT, which requires future RCTs.

Future research should be considered with larger inpatient populations in the community to evaluate whether CGM use can improve glucose management and clinical outcomes compared to standard care, while also assessing its cost-effectiveness in this setting.

Additionally, exploring optimal sensor placement and training for health care professionals may enhance usability and adherence in hospital settings. Finally, it would be important to investigate whether CGM after hospital discharge, i.e. in the community, would benefit people with comorbid diabetes and cognitive impairment, since this is such a vulnerable group for hypoglycaemic events and future hospital admissions [30].

In conclusion, CGM is highly feasible and acceptable for older hospitalised patients living with T2DM and cognitive impairment. CGM technology offers an advantage by capturing events such as asymptomatic nocturnal hypoglycaemia. Further investigation is required to understand in more detail the benefits and challenges of CGM in inpatient care for those with T2DM and cognitive impairment.

## Data Availability

All data produced in the present study are available upon reasonable request to the authors.

## Strengths and limitations of this study

- Patients and healthcare professionals blinded to CGM readings.
- Diverse recruitment across multiple health conditions and socioeconomic backgrounds.
- Small sample size and single-centre recruitment limit generalisability.
- Inclusion of acutely unwell patients may have affected study recruitment and outcomes.

## Acknowledgements

We would like to thank the participants in this study for devoting their time for this research. The views expressed are those of the author(s) and not necessarily those of the NIHR or the Department for Health and Social Care.

## Appendix

Chart 1: Participant Feedback on CGM Experience

Figure 1: Participant Recruitment Process

Figure 2: Feasibility Study Design and Procedures

Supplemental Material 1: CGM Acceptability, Feasibility and Tolerability Questionnaire *(IRAS Project ID: 344263 Version: 6.3.6 Date: 03/05/2024)*

